# The Effects of COgNitive Training in Community-dwelling Older Adults at High Risk for demENTia (CONTENT): study protocol of two double-blind, randomized, placebo-controlled trials

**DOI:** 10.1101/2025.06.27.25330394

**Authors:** Yang Pan, Mengmeng Ji, Jie Liang, Jingya Ma, Wenya Zhang, Yuling Liu, Yiwen Dai, Darui Gao, Yanyu Zhang, Wuxiang Xie, Fanfan Zheng

## Abstract

**Introduction:** Dementia contributes to the disease burden worldwide, and people with hypertension or type 2 diabetes are associated with an elevated risk of developing dementia. It is essential to prevent or delay cognitive decline in people at high risk within the community. Our trials aim to evaluate the effects of adaptive cognitive training on community-dwelling older adults with hypertension or type 2 diabetes but no dementia.

**Method and analysis:** Two multicenter, double-blind, randomized, placebo-controlled trials, named COgNitive Training in community-dwelling older adults at high risk for demENTia and with Hypertension (CONTENT-Hypertension) and COgNitive Training in community-dwelling older adults at high risk for demENTia and with Diabetes (CONTENT-Diabetes), will be conducted to investigate the effects of adaptive cognitive training on participants aged 60 years or above who have been diagnosed with hypertension or type 2 diabetes but no dementia. Each trial will enroll 120 participants. Participants will be recruited from local community in Shijingshan and Haidian District, Beijing, and allocated to either the intervention or control group using a 1:1 ratio. The intervention group will engage in 12 weeks of adaptive cognitive training, while the control group will receive 12 weeks of placebo cognitive training. A 24-week follow-up assessment will be conducted for all participants to evaluate the persistency of the effects. The primary outcome is the 12-week change in Montreal Cognitive Assessment (MoCA) Basic scores from baseline to the end of the intervention (12 weeks). Secondary outcomes include 6-week and 24-week changes in the MoCA from baseline; 6-week, 12-week, and 24-week changes in Trail Making Test-A&B (TMT-A, TMT-B), Digit Symbol Substitution Test (DSST), the World Health Organization/University of California at Los Angeles Auditory Verbal Learning Test (WHO-UCLA AVLT), and Boston naming test (BNT) scores of cognitive functions; 6-week and 12-week changes in Geriatric Depression Scale (GDS), Generalized Anxiety Disorder-7 (GAD-7), Pittsburgh sleep quality index (PSQI); and 12-week change in blood pressure (CONTENT-Hypertension) or fasting blood glucose and HbA1c (CONTENT-Diabetes) from baseline.

**Ethics and dissemination:** This study will adhere to the ethical principles outlined in the Declaration of Helsinki and comply with international standards for Good Clinical Practice (GCP). All participants will sign the informed consent at baseline. This study has been approved by the Ethics Committee of Plastic Surgery Hospital, Chinese Academy of Medical Sciences & Peking Union Medical College (approval numbers: 2023-139 and 2024-162). The findings of the trials will be disseminated through publications in peer-reviewed scientific journals and presented at academic conferences.

**Trial registration numbers:** NCT06512922 and NCT06524388.

**STRENGTHS AND LIMITATIONS OF THIS STUDY:**

- Both trials are double-blind, randomized, placebo-controlled trials to investigate the effects of adaptive cognitive training among patients with hypertension or type 2 diabetes but no dementia. The rigorous randomized control trial design can ensure the reliability and efficacy of the trial.
- Adaptive cognitive training will automatically target the most impaired cognitive domains (domain-adaptive). The difficulty of cognitive training tasks will upgrade to the next level once the participants reach a high accuracy of 80% (difficulty-adaptive), which enables targeted and efficient training.
- The use of placebo cognitive training can reduce the impact of the placebo effect on the results.
- All training will be conducted remotely, allowing participants to complete the program from their homes to increase convenience and participant compliance.

## INTRODUCTION

Dementia is a syndrome centered on the impairment of cognitive functions such as memory, attention, numerical processing, and executive control, leading to a significant deterioration in the capacity to navigate daily activities, fulfill occupational duties, and maintain social interaction.^1^ According to the Global Burden of Disease (GBD) 2019, an estimated 57.4 million people were affected by dementia globally, and this figure is anticipated to escalate dramatically 152.8 million by 2050. ^2, 3^ In 2019, the global number of disability-adjusted life years (DALYs) caused by dementia was 25.3 million, and 1.63 million people died from dementia, which makes it the seventh leading cause of death.^4^ In China, the DALYs for dementia in 2019 were 5.89 million, accounting for more than 20% of the global DALYs for dementia.^3^ There are few effective preventive and therapeutic interventions for dementia, and the drugs approved for Alzheimer’s disease cannot meet the needs of dementia treatments among the general population due to their high price, limited availability, and possible adverse effects.^5, 6^ Clearly, it remains critical to explore effective treatments for dementia.

Cumulative evidence, including our previous studies, has identified hypertension and diabetes as modifiable risk factors associated with cognitive decline.^4, 7–9^ Compared with normotensive blood pressure, hypertension demonstrates a strong correlation with more significant cognitive decline,^10, 11^ poorer cognitive function, and incident mild cognitive impairment^12^ and dementia.^13^ A worldwide study revealed that the population attributable risk of dementia due to hypertension is estimated to 15.8%.^14^ Compared with non-diabetes, patients with diabetes have 1.49 times higher risk of mild cognitive impairment and 1.43 times higher risk of dementia.^15^ A comprehensive meta-analysis from more than 2.3 million diabetic patients revealed that diabetes confers a 60% elevated risk for all-cause dementia.^16^ The latest data from the World Health Organization (WHO) indicates that the age-standardized prevalence rate of hypertension among adults aged 50 to 79 is 49% globally.^17^ The GBD study indicated that the worldwide diabetic population reached an estimated 529 million in 2021, with projections forecasting a surge to 1.31 billion by 2050.^18^ The significant accelerated cognitive decline associated with hypertension and diabetes, compounded by the escalating global incidence of hypertension and diabetes, highlights the necessity of conducting intervention studies among these patients to improve cognitive function.

Given the well-established associations of hypertension and diabetes with cognitive decline, strategies controlling blood pressure or glucose attracted increasing attention in decelerating cognitive decline or preventing dementia. However, the results were not very inspiring. Most trials to date showed antihypertensive therapy could not alleviate cognitive dysfunction,^19–21^ except for Syst-Eur,^22^ or the recent SPRINT-MIND trial.^23^ The Syst-Eur trial, which focused on older adults diagnosed with systolic hypertension, observed a 50% reduction of incident dementia with antihypertensive drug treatment, whereas the average cognitive performance assessed by Mini-mental State Examination (MMSE) was not improved. Although the SPRINT-MIND trial reported a significant effect of intensive blood pressure control (a systolic blood pressure of less than 120mmHg) on reducing the incidence of mild cognitive impairment,^23^ the possible negative impacts of intensive blood pressure control on kidney function cannot be neglected.^24^ According to the ACCORD-MIND study, intensive blood glucose therapy failed to demonstrate a cognitive benefit, and the overall risk of death was higher in the intensive blood glucose group.^25^

According to previous studies, risk factor management alone may not produce significant improvements in cognition. However, risk factor management in combination with cognitive training could significantly improve cognitive functions.^26, 27^ Despite this, one trial using multidomain intervention with cognitive training in only reasoning and memory function reported no statistically significant impact on alleviating cognitive decline,^28^ implying optimal cognitive enhancement strategies should be both comprehensive for the population and precise for each individual within the population. Therefore, we designed two double-blind, randomized controlled trials to simultaneously investigate the effects of adaptive cognitive training on older adults with hypertension or type 2 diabetes, named CONTENT-Hypertension and CONTENT-Diabetes, respectively. We hypothesized that adaptive cognitive training would be more effective in these two populations.

## METHODS AND ANALYSIS

### Study design

Both CONTENT-Hypertension and CONTENT-Diabetes are multicenter, double-blind, randomized, placebo-controlled trials targeting community-dwelling older adults aged 60 years or older with hypertension or type 2 diabetes. Participants will be recruited from the Xihuangcun community in Shijingshan District and the Peking University Hospital in Haidian District, Beijing. Eligible participants will be allocated by a 1:1 randomization ratio into either an intervention or a control group. The intervention group will engage in adaptive cognitive training and the control group will receive active cognitive training. Both training groups will be administered by using PADs with identical appearance. The training will be implemented over a 12-week period, with identical follow-up schedules for outcome assessments in both groups. Participants will be followed up at 6 weeks, 12 weeks, and 24 weeks. **Figure 1** shows the flowchart of the trials. The primary objective of our study is to estimate whether adaptive cognitive training can further effectively improve cognitive function in elderly patients with hypertension or type 2 diabetes at an early stage before the onset of dementia.

**Figure 1.**
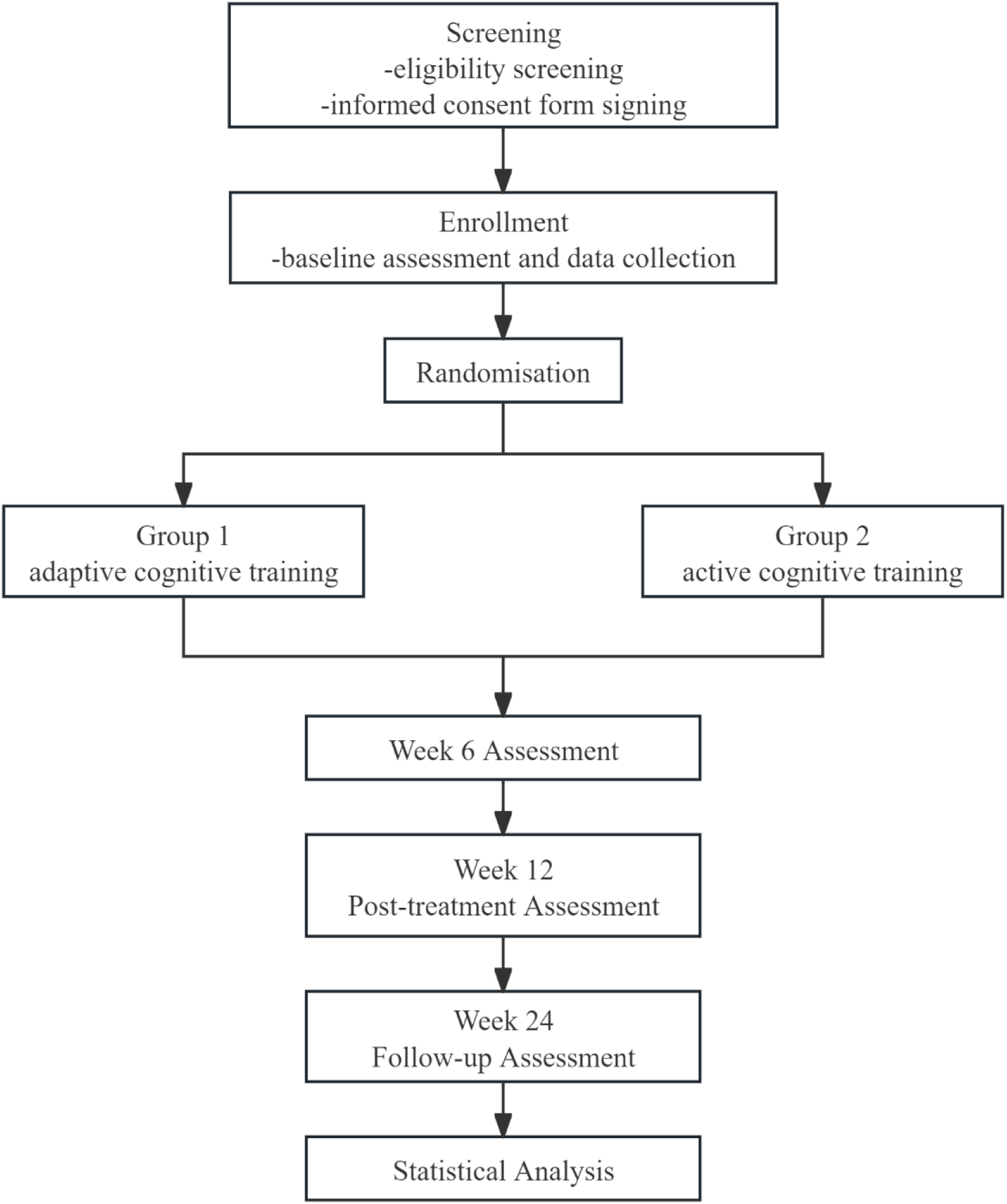
Trial flow chart.

### Participant recruitment

Individuals aged 60 years old or above, residing in the target community for a minimum of 6 months before study initiation, with a diagnosis of hypertension or type 2 diabetes, meet the criteria of these trials. Potential participants will undergo a comprehensive screening visit for eligibility and sign informed consent before entering the randomization. Detailed information on inclusion and exclusion criteria is listed below.

### Inclusion criteria

Inclusion criteria included the following: (1) Participants were clinically diagnosed with hypertension, systolic blood pressure (SBP) ranged 140-180 mmHg on no medication or no limitation of SBP on 1 or more medications (CONTENT-Hypertension); participants were clinically diagnosed with type 2 diabetes, fasting blood glucose>6.1mmol/L (CONTENT-Diabetes); and do not intent to change the treatment regimen for the disease during the expected duration of the trial; (2) Literate in Han Chinese, age ≥60 years; (3) CAIDE risk score ≥ 6,^29–32^ but no dementia; (4) Living in the community with a minimum 6-month residency prior to the trial and maintaining residence in the community for at least 6 months; (5) Signed informed consent.

### Exclusion criteria

Exclusion criteria included the following: (1) Could not complete neuropsychological tests; (2) Suffering other disease or medication use might affect cognitive function, such as a new stroke within 6 months before baseline or use tranquilizers; (3) Unable to use PADs or unable to complete the training; (4) Unacceptable or refuse to participate in cognitive training; (5) Living in the same family with a subject who has been randomly assigned.

### Randomization and blinding

Following the completion of informed consent procedures and baseline assessments, eligible participants will undergo randomization by an independent biostatistician, using SAS version 9.4 (SAS Institute, Cary, NC). A block randomization with variable block sizes will be implemented to allocate participants to either intervention or control group in a 1:1 ratio. If participants failed to initiate the training protocol within 7 days after randomization, they were withdrawn from the trial and their randomization number was reassigned to a new eligible participant. Each randomization number will be permitted a maximum of two reassignments. To uphold the concealment of group allocation, the allocation for each participant will be directly imported into the intervention system. The intervention and control group will use PADs with identical appearances to deliver cognitive training throughout the study, so both participants and researchers will be masked to the intervention allocation. Additionally, researchers responsible for outcome evaluations and statistical analyses will also be masked.

### Interventions

Participants allocated to the intervention group will engage in a 12-week, computerized, active cognitive training. This comprehensive training program is designed to target a broad spectrum of neuropsychological functions, including processing speed, attention, perception, long-term and working memory, thinking, executive control, inductive and reasoning skill, and linguistic abilities. Cognitive training tasks have been developed under these domains based on classical intervention paradigms, such as n-back, Corsi block-tapping, go/no-go, flanker, etc. All these tasks are designed to enable a game-like interactive experience to improve adherence. A self-adaptive algorithm is embedded in the cognitive training application, which will help deliver suitable tasks at the appropriate difficulty level for each participant according to their profile and real-time performance. Participants will be required to complete daily 30-minute training sessions for a minimum of five days weekly. The difficulty level of each task will automatically escalate once consistent high accuracy is demonstrated (e.g., exceeding 80% correct responses).

The active control group will engage in a comparable training regimen, completing 5 tasks daily for a total of 30 minutes, with a minimum frequency of 5 days per week. In contrast to the intervention group, these tasks will maintain a predetermined, constant difficulty level throughout the study duration.

All participants will engage in the cognitive training at home and be monitored by an independent researcher over the Internet. The researcher will contact the participant once he/she did not fulfill the training during the past week to identify the reasons and ensure the adherence.

### Baseline data collection

Data will be collected from all participants. Baseline data collection includes a questionnaire interview on sociodemographic characteristics (age, sex, employment status, educational level, marital status, household income, and health insurance), lifestyle and health behaviors (tobacco use, alcohol consumption, physical activity, and dietary habits), clinical history (depression, stroke, myocardial infarction, angina, atrial fibrillation, heart failure, chronic kidney disease, chronic obstructive pulmonary disease, and cancer), medication profile (antidepressant drugs, antihypertensive drugs, antidiabetic drugs, and lipid-lowering drugs), depressive symptoms by using GDS, daily and social activity function by ADL and FAQ, anxious symptoms by GAD-7, sleep quality by PSQI, and cognitive function assessed by using the MoCA, TMT-A, TMT-B, DSST, the WHO-UCLA AVLT, and BNT. Physical examinations include blood pressure, height, weight, pulse rate, and waist circumference. Laboratory assessments include fasting blood tests (creatinine, uric acid, alanine aminotransferase, fasting blood glucose, glycated hemoglobin (HbA_1c_), total cholesterol, low-density lipoprotein cholesterol, high-density lipoprotein cholesterol, and triglycerides) and apolipoprotein E4 (APOE4) genotype.

### Methods of physical measurements

All physical measurements will be conducted by professionally trained nurses following Standard Operating Procedure (SOP). Height and weight measurements will be completed using a standard integrated height and weight measuring device, and participants will be asked to remove shoes and hats for height measurements and excess clothing for weight measurements. Body Mass Index (BMI) is calculated by weight (kg) divided by the square of height (m^2^). Waist circumference will be measured by using a uniform tape measure. Systolic blood pressure, diastolic blood pressure (DBP), and pulse rate will be examined using calibrated Omron J-710 automated monitor. Blood pressure will be measured three times to ensure accuracy, with a mandatory rest period of at least 1minute apart.^33^ The final blood pressure value is the average of the last two blood pressure.^34^

### Blood samples collection

Participants’ blood samples will be obtained by nurses from community hospitals after a minimum 8-hour abstention from food and fluid intake. Following centrifugation, the serum samples will undergo stringent temperature-controlled processing and transportation through an uninterrupted cold chain. Laboratory assessment will be conducted in Lawke Health Laboratory and BGI Genomics Company in Beijing.

### Questionnaire interview on demographic characteristics and cognitive function

In this study, trained researchers will administer structured questionnaires in person to collect sociodemographic information, lifestyle information, disease history, medication information, depressive symptoms, daily and social activity function, anxious symptoms, sleep quality, and cognitive function assessment from all participants. Before the initiation of the study, all researchers will attend a comprehensive training program, and only those who pass the examination after training will be eligible to conduct the baseline and follow-up investigations.

### Follow-up schedules

Follow-up visits are planned at 6 weeks, 12 weeks (the end of the intervention), and 24 weeks (12 weeks after the end of intervention). Assessment methods identical to those at baseline will be employed for follow-up questionnaire interviews and physical examinations.

### Data management

We will employ H6space (https://h6world.cn) to streamline data collection and central management throughout the study duration. H6space is a free online Electronic Data Capture (EDC) system developed by Peking University for collecting, managing, analyzing, and sharing whole-life clinical and biological sample data. In our study, a unique study ID will be assigned to identify each participant’s data. Data integrity and quality assurance will be maintained through a comprehensive validation process.

This includes automated h6space validation checks complemented by meticulous manual verification procedures. To ensure confidentiality and compliance with data protection regulations, access to study information is strictly limited to authorized research personnel. All physical documentation, including Case Report Forms (CRFs), will be securely stored in locked cabinets within designated restricted areas. Trial documentation and data will be preserved in secure archives for a minimum of 5 years following study completion.

### Outcomes

#### Primary outcome

The primary outcome is the change in MoCA scores from baseline to the end of the intervention (12 weeks).

#### Secondary outcomes

The secondary outcomes include 6-week and 24-week changes in the MoCA from baseline; 6-week, 12-week, and 24-week changes in TMT-A, TMT-B, DSST, the WHO-UCLA AVLT, and BNT scores of cognitive functions; 6-week and 12-week changes in GDS, GAD-7, PSQI, and 12-week change in blood pressure or fasting blood glucose and HbA1c from baseline.

#### Sample size

According to our previous findings from our Beijing Research on Ageing and Vessel (BRAVE) cohort study, the mean ± standard deviation (SD) of MoCA scores was 23.6 ± 3.5 among community-dwelling participants aged ≥ 60 years with a CAIDE risk score ≥ 6.^35^ Our pilot trial also found that the mean MoCA scores of baseline was 21.3 ± 3.5 (N=20), and a 4-week cognitive training improved MoCA scores from 21.3 to 25.6. Based on previous evidence, we assume that the SD of MoCA scores of our participants will be 3.5, and our 12-week adaptive cognitive training will lead to an absolute increase of 2.0 in MoCA scores compared with the active control group (a change of 0.5 SD or over in a health-related indicator is deemed to have clinical significance^36^). The current study was powered to detect a difference of 2.0 in MoCA between randomization groups, with 80% power and a type I error rate of 5%, a sample of 50 participants per group will be required. Considering a 15% attrition rate, this resulted in a required total sample size of 120 per trial (60 participants per group).

## Statistical analysis

Primary analyses will be performed by using the modified intention-to-treat approach, which will exclude individuals who do not receive any cognitive training and do not attend any follow-up visits for cognitive function. The results will be presented as frequency and proportion for categorical variables and means ± SDs for normally distributed continuous variables. To investigate the effects of adaptive cognitive training on outcomes, linear mixed effect models nested within individuals will be used. Time will be designated as the repeated variable. The fixed effects will comprise the group, time, and group×time interaction, and group×time interaction term indicates differential change by group from baseline to end of the trial (or other time-point).

Per-protocol analyses will be conducted as a sensitive analysis. This subset will include participants who receive 80% designated cognitive training and attended the 6-week and 12-week follow-up assessment for cognitive function. Subgroup analyses will be conducted to explore the potential modifiers of active cognitive training effect, such as age, sex, baseline cognitive function, educational level, marital status, and clinical history.

All statistical analyses will be conducted using SAS version 9.4 (SAS Institute Inc., Cary, NC, USA). All analyses will be two-sided; an alpha value of 0.05 will be considered as the threshold for statistical significance.

### Patient and public involvement

This study is conceptualized, designed, and will be executed without involvement of patients or members of the public in implementation, reporting, or dissemination of this trial.

### Ethics and dissemination

This study adheres to the Declaration of Helsinki and guidelines of good clinical practice. All participants will sign informed consent prior to any assessment and data collection. Participant data will be managed through a secure in the EDC system. Access to the system is strictly controlled and limited to authorized study personnel, each assigned specific permission levels commensurate with their roles and responsibilities. To maintain participant anonymity and comply with data protection regulations, all statistical analyses will be conducted using de-identified data.

The results of this study will be published in international peer-reviewed journals and disseminated through academic conferences.

### Quality control

All researchers must attend the standardized training, including study protocol, informed consent, CRFs, SOP for data collection and management, and collection methods of blood samples. Additionally, regular team meetings will be held regularly to discuss operational challenges and maintain adherence to the established protocol and SOP throughout the study duration. The research team will convene regularly to discuss operational challenges and maintain adherence to the established protocol and predefined procedural guidelines throughout the study duration.

Blood samples will be tested in Lawke Health Laboratory and BGI Genomics Company located in Beijing. Laboratory personnel conducting biochemical and genetic analyses will remain unaware of participant’s randomization allocation. Adherence monitoring will be conducted to ensure completion of cognitive training and overall engagement with the trial.

### Current status

This trial is in enrollment, and the first participant was enrolled in October 29, 2024.

## Discussion

Considering the disease burden and progress of clinical treatment for dementia, preventive intervention remains one of the most important strategies,^37, 38^ especially for older adults with hypertension or type 2 diabetes demonstrating early signs of cognitive decline prior to the onset of dementia.^7, 39^ To the best of our knowledge, CONTENT-Hypertension and CONTENT-Diabetes trials will be the first study to simultaneously investigate the effects of adaptive cognitive training among older patients with hypertension or type 2 diabetes with no dementia.

Cognitive training combined risk factors management could significantly improve cognitive functions.^26, 27^ Based on the classical cognitive training paradigm, adaptive cognitive training develops interactive game tasks to enhance multidomain cognitive function. The training materials are carefully selected from real-life scenarios, and timely encouragement and feedback are provided throughout the process. As a result, it not only effectively alleviates participants’ cognitive function but also increases their adherence to the training program.^40^ As older adults increasingly use and rely on electronic devices, home cognitive training with electronic devices becomes more appealing, and easier to extend the intervention to more people.^37^ Previous trials have demonstrated the advantages of adaptive cognitive training over general cognitive training in patients with mild cognitive impairment, neuropsychiatric symptoms, and multiple sclerosis.^41, 42^ Regarding diabetes, only two studies comparing adaptive and general cognitive training, but neither showed a significant advantage of adaptive cognitive training.^43, 44^ These differences may be due to differences in participants, sample size, attrition rate, cognitive training paradigm, and the duration of the intervention.

Study design and quality control measures ensure the reliability of the study results. The intervention of 12 weeks and follow-up of 24 weeks will provide a unique opportunity to indicate the enduring effect of cognitive training on the population with hypertension and type 2 diabetes. To mitigate potential placebo effect on the results, we select active cognitive training as a control group. Furthermore, we will conduct two parallel randomized controlled trials for hypertension and type 2 diabetes, focusing on community-dwelling older adults at high risk of dementia, to enhance the generalizability of our results.

In conclusion, this study will further investigate the effect of adaptive cognitive training for community-dwelling older patients with hypertension and diabetes. It aims to provide new evidence concerning the effect of cognitive training among older adults at high risk for dementia and to analysis whether adaptive cognitive training can yield additional benefits. Findings of our trials may shed light on the development of preventive strategies for dementia.

## Contributions

FZ and WX conceived the original idea for the trial and are responsible for the overall content as guarantors. YP and MJ developed the trial design and wrote the manuscript. FZ and WX wrote the statistical analysis plan and revised the manuscript. JL, JM, WZ, YL, YD, DG, and YZ has been part of the trial design. All authors read and approved the manuscript.

## Data Availability

Not applicable.

## References

1. WHO. Dementia. 2023. Vol 2024.

2. Estimation of the global prevalence of dementia in 2019 and forecasted prevalence in 2050: an analysis for the Global Burden of Disease Study 2019. Lancet Public Health. 2022;7(2): e105–e125.

3. Global burden of 369 diseases and injuries in 204 countries and territories, 1990-2019: a systematic analysis for the Global Burden of Disease Study 2019. Lancet. 2020;396(10258):1204–1222.

4. Jia L, Du Y, Chu L, et al. Prevalence, risk factors, and management of dementia and mild cognitive impairment in adults aged 60 years or older in China: a cross-sectional study. Lancet Public Health. 2020;5(12): e661–e671.

5. Jucker M, Walker LC. Alzheimer’s disease: From immunotherapy to immunoprevention. Cell. 2023;186(20):4260–4270.

6. Whitehouse P, Gandy S, Saini V, et al. Making the Case for Accelerated Withdrawal of Aducanumab. J Alzheimers Dis. 2022;87(3):1003–1007.

7. Livingston G, Huntley J, Sommerlad A, et al. Dementia prevention, intervention, and care: 2020 report of the Lancet Commission. Lancet. 2020;396(10248):413–446.

8. Li C, Zhu Y, Ma Y, Hua R, Zhong B, Xie W. Association of Cumulative Blood Pressure With Cognitive Decline, Dementia, and Mortality. J Am Coll Cardiol. 2022;79(14):1321–1335.

9. Zheng F, Yan L, Yang Z, Zhong B, Xie W. HbA(1c), diabetes and cognitive decline: the English Longitudinal Study of Ageing. Diabetologia. 2018;61(4):839–848.

10. Gottesman RF, Schneider AL, Albert M, et al. Midlife hypertension and 20-year cognitive change: the atherosclerosis risk in communities neurocognitive study. JAMA Neurol. 2014;71(10):1218–1227.

11. Swan GE, DeCarli C, Miller BL, et al. Association of midlife blood pressure to late-life cognitive decline and brain morphology. Neurology. 1998;51(4):986–993.

12. Kivipelto M, Helkala EL, Hanninen T, et al. Midlife vascular risk factors and late-life mild cognitive impairment: A population-based study. Neurology. 2001;56(12):1683–1689.

13. Gottesman RF, Albert MS, Alonso A, et al. Associations Between Midlife Vascular Risk Factors and 25-Year Incident Dementia in the Atherosclerosis Risk in Communities (ARIC) Cohort. JAMA Neurol. 2017;74(10):1246–1254.

14. Mulligan MD, Murphy R, Reddin C, et al. Population attributable fraction of hypertension for dementia: global, regional, and national estimates for 186 countries. EClinicalMedicine. 2023; 60:102012.

15. Xue M, Xu W, Ou YN, et al. Diabetes mellitus and risks of cognitive impairment and dementia: A systematic review and meta-analysis of 144 prospective studies. Ageing Res Rev. 2019; 55:100944.

16. Chatterjee S, Peters SA, Woodward M, et al. Type 2 Diabetes as a Risk Factor for Dementia in Women Compared With Men: A Pooled Analysis of 2.3 Million People Comprising More Than 100,000 Cases of Dementia. Diabetes Care. 2016;39(2):300–307.

17. WHO. Global report on hypertension:The race against a silent killer; 2023.

18. Global, regional, and national burden of diabetes from 1990 to 2021, with projections of prevalence to 2050: a systematic analysis for the Global Burden of Disease Study 2021. Lancet. 2023;402(10397):203–234.

19. Zonneveld TP, Richard E, Vergouwen MD, et al. Blood pressure-lowering treatment for preventing recurrent stroke, major vascular events, and dementia in patients with a history of stroke or transient ischaemic attack. Cochrane Database Syst Rev. 2018;7(7): D7858.

20. Ihle-Hansen H, Thommessen B, Fagerland MW, et al. Blood pressure control to prevent decline in cognition after stroke. Vasc Health Risk Manag. 2015; 11:311–316.

21. Bath PM, Scutt P, Blackburn DJ, et al. Intensive versus Guideline Blood Pressure and Lipid Lowering in Patients with Previous Stroke: Main Results from the Pilot’Prevention of Decline in Cognition after Stroke Trial’ (PODCAST) Randomised Controlled Trial. PLoS One. 2017;12(1): e164608.

22. Forette F, Seux ML, Staessen JA, et al. Prevention of dementia in randomised double-blind placebo-controlled Systolic Hypertension in Europe (Syst-Eur) trial. Lancet. 1998;352(9137):1347-1351.

23. Williamson JD, Pajewski NM, Auchus AP, et al. Effect of Intensive vs Standard Blood Pressure Control on Probable Dementia: A Randomized Clinical Trial. JAMA. 2019;321(6):553–561.

24. Kurella TM, Gaussoin SA, Pajewski NM, et al. Kidney Disease, Intensive Hypertension Treatment, and Risk for Dementia and Mild Cognitive Impairment: The Systolic Blood Pressure Intervention Trial. J Am Soc Nephrol. 2020;31(9):2122–2132.

25. Murray AM, Hsu FC, Williamson JD, et al. ACCORDION MIND: results of the observational extension of the ACCORD MIND randomised trial. Diabetologia. 2017;60(1):69–80.

26. Ngandu T, Lehtisalo J, Solomon A, et al. A 2 year multidomain intervention of diet, exercise, cognitive training, and vascular risk monitoring versus control to prevent cognitive decline in at-risk elderly people (FINGER): a randomised controlled trial. Lancet. 2015;385(9984):2255-2263.

27. Lee KS, Lee Y, Back JH, et al. Effects of a multidomain lifestyle modification on cognitive function in older adults: an eighteen-month community-based cluster randomized controlled trial. Psychother Psychosom. 2014;83(5):270–278.

28. Andrieu S, Guyonnet S, Coley N, et al. Effect of long-term omega 3 polyunsaturated fatty acid supplementation with or without multidomain intervention on cognitive function in elderly adults with memory complaints (MAPT): a randomised, placebo-controlled trial. Lancet Neurol. 2017;16(5):377–389.

29. Sindi S, Calov E, Fokkens J, et al. The CAIDE Dementia Risk Score App: The development of an evidence-based mobile application to predict the risk of dementia. Alzheimers Dement (Amst). 2015;1(3):328–333.

30. Kivipelto M, Ngandu T, Laatikainen T, Winblad B, Soininen H, Tuomilehto J. Risk score for the prediction of dementia risk in 20 years among middle aged people: a longitudinal, population-based study. Lancet Neurol. 2006;5(9):735–741.

31. Kaffashian S, Dugravot A, Elbaz A, et al. Predicting cognitive decline: a dementia risk score vs. the Framingham vascular risk scores. Neurology. 2013;80(14):1300–1306.

32. Ngandu T, Lehtisalo J, Solomon A, et al. A 2 year multidomain intervention of diet, exercise, cognitive training, and vascular risk monitoring versus control to prevent cognitive decline in at-risk elderly people (FINGER): a randomised controlled trial. Lancet. 2015;385(9984):2255-2263.

33. Williams B, Mancia G, Spiering W, et al. 2018 ESC/ESH Guidelines for the management of arterial hypertension. Eur Heart J. 2018;39(33):3021-3104.

34. Beaney T, Schutte AE, Tomaszewski M, et al. May Measurement Month 2017: an analysis of blood pressure screening results worldwide. Lancet Glob Health. 2018;6(7): e736–e743.

35. Lu Y, Zhu Y, Ma Y, et al. Association of subclinical atherosclerosis and cognitive decline: a community-based cross-sectional study. BMJ Open. 2022;12(5): e59024.

36. Norman GR, Sloan JA, Wyrwich KW. Interpretation of changes in health-related quality of life: the remarkable universality of half a standard deviation. Med Care. 2003;41(5):582–592.

37. Rosenberg A, Mangialasche F, Ngandu T, Solomon A, Kivipelto M. Multidomain Interventions to Prevent Cognitive Impairment, Alzheimer’s Disease, and Dementia: From FINGER to World-Wide FINGERS. J Prev Alzheimers Dis. 2020;7(1):29–36.

38. Grande G, Qiu C, Fratiglioni L. Prevention of dementia in an ageing world: Evidence and biological rationale. Ageing Res Rev. 2020; 64:101045.

39. Xu W, Tan L, Wang HF, et al. Meta-analysis of modifiable risk factors for Alzheimer’s disease. J Neurol Neurosurg Psychiatry. 2015;86(12):1299–1306.

40. Lee HK, Kent JD, Wendel C, et al. Home-Based, Adaptive Cognitive Training for Cognitively Normal Older adults: Initial Efficacy Trial. J Gerontol B Psychol Sci Soc Sci. 2020;75(6):1144–1154.

41. Bahar-Fuchs A, Webb S, Bartsch L, et al. Tailored and Adaptive Computerized Cognitive Training in Older Adults at Risk for Dementia: A Randomized Controlled Trial. J Alzheimers Dis. 2017;60(3):889–911.

42. Pedulla L, Brichetto G, Tacchino A, et al. Adaptive vs. non-adaptive cognitive training by means of a personalized App: a randomized trial in people with multiple sclerosis. J Neuroeng Rehabil. 2016;13(1):88.

43. Bahar-Fuchs A, Barendse M, Bloom R, et al. Computerized Cognitive Training for Older Adults at Higher Dementia Risk due to Diabetes: Findings From a Randomized Controlled Trial. J Gerontol A Biol Sci Med Sci. 2020;75(4):747–754.

44. Silverman JM, Zhu CW, Schmeidler J, et al. Does computerized cognitive training improve diabetes self-management and cognition? A randomized control trial of middle-aged and older veterans with type 2 diabetes. Diabetes Res Clin Pract. 2023; 195:110149.

